# Isolation and Characterization of Human Adipocyte-derived Extracellular Vesicles Using Filtration and Ultracentrifugation

**DOI:** 10.1101/2020.12.03.20243089

**Authors:** Naveed Akbar, Katherine E. Pinnick, Daan Paget, Robin P. Choudhury

## Abstract

Extracellular vesicles (EVs) are lipid enclosed envelopes that carry biologically active material such as proteins, RNA, metabolites and lipids. EVs can modulate the cellular status of other cells locally in tissue microenvironments or through liberation into peripheral blood. Adipocyte-derived EVs are elevated in the peripheral blood and show alterations in their cargo (RNA and protein) during metabolic disturbances including, obesity and diabetes. Adipocyte-derived EVs can regulate the cellular status of neighboring vascular cells, such as endothelial cells and adipose tissue resident macrophages to promote adipose tissue inflammation. Investigating alterations in adipocyte-derived EVs in vivo is complex because EVs derived from peripheral blood are highly heterogenous and contain EVs from other sources, namely platelets, endothelial cells, erythrocytes and muscle. Therefore, the culture of human adipocytes provides a model system for the study of adipocyte derived EVs. Here, we provide a detailed protocol for the extraction of total small EVs from cell culture media of human gluteal and abdominal adipocytes using filtration and ultracentrifugation. We further demonstrate the use of Nanoparticle Tracking Analysis (NTA) for quantification of EV size and concentration and show the presence of EV-protein tumor susceptibility gene 101 (TSG101) in the gluteal and abdominal adipocyte derived-EVs. Isolated EVs from this protocol can be used for downstream analysis including, transmission electron microscopy, proteomics, metabolomics, small RNA-sequencing, microarray and utilized in functional in vitro/in vivo studies.

**SUMMARY:** We describe the isolation of human adipocyte-derived extracellular vesicles (EVs) from gluteal and abdominal adipose tissue using filtration and ultracentrifugation. We characterize the isolated adipocyte-derived EVs by determining their size and concentration by Nanoparticle Tracking Analysis and by western blotting for the presence of EV-protein tumor susceptibility gene 101 (TSG101).

## INTRODUCTION

Extracellular vesicles (EVs) are lipid enclosed envelopes that carry biologically active material such as proteins, microRNAs, metabolites and lipids. The term EVs denotes various subpopulations, which include exosomes, microvesicles (microparticles/ectosomes) and apoptotic bodies^1^. EVs may serve as biomarkers because they are implicated in pathological signaling and released into bio fluids including blood and urine. EVs can modulate the cellular status of other cells locally in tissue microenvironments or through liberation into peripheral blood^2^. EVs bear features of their parent cell but differentiation of each sub population is primarily based on EVs size and protein content such as EVs markers, including the presence of tetraspanins (CD9, CD63 and CD81), tumor susceptibility gene 101 (TSG101) and ALG-2- interacting protein X (ALIX). These protein markers are representative of the endosomal origin (CD9, CD63 and CD81) for exosomes, which are generated inside multi-vesicular bodies or represent proteins associated with budding or blebbing directly from the plasma membrane for microvesicles. However, there is a significant overlap between these subpopulations and it is difficult to distinguish individual sub-populations in complex bio fluids such as plasma, serum or urine.

Metabolic disturbances, including obesity, insulin resistance, and perturbations in extracellular glucose, oxygen and inflammation can alter the size and concentration of EVs and their cargo. Adipocyte-derived EVs carry Perilipin A, adiponectin and show alterations in their protein and RNA cargo during obesity and diabetes^3-6^. Adipocyte-derived EVs regulate the cellular status of neighboring vascular endothelial cells^7^ and adipose tissue resident macrophages to promote adipose tissue inflammation and insulin resistance^8-11^. Investigating alterations in adipocyte-derived EVs in vivo is complex because EV populations derived from complex biofluids such as plasma, serum or urine contain EVs from multiple sources, such as platelets, endothelial cells, erythrocytes and muscle, which are implicated in the pathogenesis of metabolic dysfunction and disease.

The culture and in vitro differentiation of human preadipocytes, therefore, provides a model system for the study of adipocyte derived EVs. Here, we provide a detailed protocol for the extraction of total small EVs from cell culture media of human adipocytes using syringe filtration and ultracentrifugation. Ultracentrifugation remains a popular method of isolation for EVs because it is easily accessible and requires little prior specialist knowledge. However, other methods such as precipitation, size exclusion chromatography and immunoaffinity capture using tetraspanins enable EV isolation from a range of biofluids including plasma, serum, urine and conditioned cell culture media. Each method, including the ultracentrifugation protocol described here produce EV preparations of varying purity because the methods can co-isolate soluble proteins and lipoproteins, which may mask as EVs. Combining this ultrancentrifugation protocol with other methods such as density centrifugation, size, exclusion chromatography and immune-affinity capture dramatically increases the purity of isolated EVs. But similar to ultracentrifugation, these other methods do not allow the capture of independent sub- populations of EVs from complex samples such as blood, plasma and urine. Therefore, cultures of selected cell populations remain one of the most robust methods for generating high yields of cell specific EVs. Each EV method has a number of caveats and the choice of method can impact the types of EVs isolated and their concentrations, which may bias downstream mechanistic investigations into cellular and tissue signaling and determination of EV-cargo for diagnostic studies; these methodological issues of EV isolation are discussed elsewhere and in the limitation section below^4,12^. Here, we describe the isolation of human adipocyte derived EVs using filtration and ultracentrifugation. We further demonstrate the use of Nanoparticle Tracking Analysis (NTA) for quantification of EV size and concentration and show the presence of EV-protein tumor susceptibility gene 101 (TSG101) in our human adipocyte derived EVs. Isolated EVs from this protocol can be used for downstream analysis, including transmission electron microscopy, proteomics, metabolomics, small RNA-sequencing, microarray and utilized in functional in vitro/in vivo studies.

## PROTOCOL

All methods were approved by the institutional ethics review board at the University of Oxford. Adipose tissue was obtained by needle biopsy under local anesthetic from healthy volunteers.

### 1. Preparation of cell culture medium and buffers

1.1. Prepare a collagenase digestion buffer by dissolving collagenase H (1 mg/mL) in Hanks Balanced Salt Solution (HBSS) (without calcium chloride and without magnesium chloride) and sterile filter using a 0.2 µm pore syringe filter.
1.1.1. Prepare the collagenase digestion buffer no more than 10 minutes before use.
1.2. Prepare growth medium (GM) as follows: Dulbecco’s Modified Eagle’s Medium/Ham’s nutrient mixture F12 (DMEM/F12) supplemented with 10% fetal bovine serum (FBS), 100 units/mL penicillin, 100 µg/mL streptomycin, 2 mM L-glutamine and 0.5 ng/mL fibroblast growth factor (FGF).
1.3. Prepare the basic differentiation medium (basic DM) as follows: DMEM/F12 supplemented with 100 units/mL penicillin, 100 µg/mL streptomycin, 2 mM L-glutamine, 17 μM pantothenate, 100 nM human insulin, 10 nM triiodo-L-thyronine, 33 μM biotin, 10 μg/mL transferrin and 1 μM dexamethasone.
1.4. Prepare complete differentiation medium (complete DM) by supplementing the basic DM from step 1.3 with 0.25 mM 3-isobutyl-1-methylxanthine and 4 μM troglitazone.
1.5. Prepare 10 mM fatty acid stock solutions complexed to 10% essentially fatty acid free bovine serum albumin (BSA) as follows.
1.5.1. Dissolve 16 g of BSA in 160 mL of DMEM/F12 medium and warm to 37 °C. In three separate 50 mL conical tubes, weigh out 150 mg of sodium oleate, 139 mg of sodium palmitate and 151 mg of sodium linoleate. Add 50 mL of the warmed BSA solution to each tube and mix well by repeated vortexing.
1.5.2. Return the oleate and linoleate tubes to the 37 °C water bath for 15 minutes. Mix by vortexing until completely dissolved.
1.5.3. For the palmitate solution place the tube in a 65 °C water bath for 2-3 minutes. Mix vigorously by vortexing.
1.5.4. Repeat step 1.5.3. until fully dissolved – approximately 30 minutes. NOTE: Some small particles may still be visible.
1.5.5. Sterile filter the fatty acid solutions using a 0.2 µm pore syringe filter.
1.5.6. Confirm the non-esterified fatty acids concentration of each stock solution using an appropriate assay. NOTE: Fatty acid stocks should be 10 mM (± 10%) to give a molar ratio of 6:1 between fatty acid and BSA. If the concentration is outside this range the stock solution should be remade. Stock solutions can be aliquoted and stored at -30 °C.

### 2. Digestion of human adipose tissue biopsies

2.1. Slowly strain the contents of the biopsy syringe through a 200 µm sterile cell strainer attached to a 50 mL conical tube so that the adipose tissue is collected in the strainer.
2.2. Transfer the strainer to a new 50 mL conical tube and wash the adipose tissue three times with 10 mL of HBSS. NOTE: It may be necessary to remove blood clots and fibrous tissue with surgical scissors or to repeat the HBSS wash to remove excess red blood cells.
2.3. Weigh the washed tissue. NOTE: We typically obtain 400-800 mg of tissue by needle biopsy.
2.4. Put the collagenase digestion buffer in a sterile 50 mL conical tube and add the washed tissue. Surgical scissors can be used at this step to mince the tissue into equal sized pieces. NOTE: Use 5 mL of buffer per 0.5 g tissue. For larger tissue, samples can be minced in a Petri dish.
2.5. Place the tube into a 37 °C shaking water bath and incubate for 35 - 40 minutes. NOTE: Following successful digestion the solution should appear milky. If small pieces of tissue are still visible shake by hand for a further 10 - 20 s.

### 3. Isolation of preadipocytes

3.1. To pellet the preadipocyte fraction centrifuge at 1000 x *g* for 5 minutes.
3.2. Aspirate and discard the floating adipocyte layer and supernatant leaving approximately 1 mL of HBSS covering the cell pellet.
3.3. Resuspend the pellet in 5 mL of HBSS and pass the cell suspension through a 250 μm pore size mesh followed by a 100 μm mesh to remove any undigested material. Collect the cell suspension in a 15 mL conical tube.
3.4. Centrifuge at 1000 x *g* for 5 minutes. NOTE: After this step, the preadipocyte fraction can be treated with a red blood cell lysis solution if red blood cell contamination is an issue.
3.5. Aspirate and discard the supernatant.
3.6. Resuspend the cell pellet in 5 mL of GM (step 1.2) and seed into a 25 cm^2^ adherent tissue culture flask. Place the flask in a cell culture incubator (5% CO_2_, 37 °C).

### 4. Maintenance of preadipocyte cultures

4.1. Replace the GM every 2 days while the cells are proliferating NOTE: Cell proliferation rates may vary between donors.
4.1.1. When the cells reach approximately 80% confluence transfer them to a 75 cm^2^ adherent tissue culture flask. Remove the GM and wash the cells with phosphate buffered saline (PBS).
4.1.2. Discard the PBS and add 0.5 mL of highly purified cell dissociation enzymes to disassociate the attached cells. Incubate at 37 °C for 5 minutes.
4.1.3. Tap the flask sharply to release the cells and add 5 mL of GM. Collect the cell suspension in a 15 mL conical tube.
4.2. Centrifuge at 1000 x *g* for 5 minutes
4.3. Aspirate and discard the supernatant. Resuspend the cell pellet in 5 mL of GM and transfer to a 75 cm^2^ flask. Top up the GM to a final volume of 12 mL and replace the flask in the cell culture incubator (5% CO_2_, 37 °C). NOTE: To maintain the cells continue to change the GM every 2-3 days. When the cells reach 80% confluence repeat steps 4.2-4.3 and divide the cells 1:3 into new 75 cm^2^ flasks. This can be repeated for several passages until sufficient number of cells have been generated for the experimental set up. We would not recommend more than 10 passages as proliferation and differentiation capacity of these preadipocytes declines.

### 5. Seeding preadipocytes for adipogenic differentiation

5.1. Count the cells using a hemocytometer and seed 200,000 cells into multiple wells of a 6 well plate in 2 mL of GM. Alternatively seed 3.5 million cells per T175 cm^2^ flask in 22 mL of GM media.
5.2. Allow the cells to proliferate for a further 2--4 days until they reach full confluence, changing the GM on the second day of culture.
5.3. To begin adipogenic differentiation remove the GM and replace with complete DM for 4 days (2 mL of complete DM per well of a 6 well plate or 22 mL for a T175 cm^2^ flask. NOTE: Replace with fresh complete DM on day 2.
5.4. On day 4 remove the complete DM and replace with 2 mL basic DM supplemented with 22.5 μM oleate, 15 μM palmitate and 12.5 μM linoleate to give a total fatty acid concentration of 50 μM per well of a 6 well plate or 22 mL for a T175 cm^2^ flask. Replace with media every 2 days for a further 10 days. NOTE: From day 7 onwards lipid droplets should be visible in the differentiating preadipocytes.
5.5. On culture day 14 collect the media from the cells for isolation of adipocyte-derived EVs

### 6. Preparation of cell culture supernatant for extracellular vesicle isolation or storage and future extracellular vesicle isolation

6.1. Remove cell culture supernatants from each 6 well plate and combine. Add to a 15 mL tube or remove all the cell culture supernatant from a T175 cm^2^ flask and add to a 50 mL tube.
6.2. Centrifuge for 10 minutes are 4 °C at 1000 x *g*.
6.3. Decant supernatant in to a new clean 15 mL or 50 mL tube by pouring, respectively.
6.4. Remove the barrel of a 10 mL syringe and attach 0.45 µm syringe filter.
6.5. Pour in the supernatant to the syringe reservoir and apply gentle pressure at the syringe barrel opening with a thumb or palm until the cell culture supernatant freely passes through the filter. NOTE: The speed of this filtration step can vary depending on the type of syringe filter.
6.6. Collect the filtrate in a clean 50 mL tube. NOTE: If required the conditioned filtered media can be stored at this point at -80 °C for several weeks. When required defrost the cell culture supernatants at 4 °C and centrifuge for 10 minutes are 4 °C at 1000 x *g*. Decant supernatant into a clean tube by pouring before continuing with the EV isolations.
6.7. Collect the filtrate in a clean 50 mL tube.

### 7. Isolation of extracellular vesicles

7.1. Label a 13 mL ultracentrifugation tube by drawing a circle at the bottom of the tube, where the expected EV pellet will form and mark a line around the neck of the tube for orientation in the ultracentrifugation tube rotor. Label the tube with a sample identifier.
7.2. Place tube in tube holder.
7.3. Attach a 16G needle to a 10 mL syringe, and remove the syringe barrel.
7.4. Remove the protective cover from needle and insert the needle into the neck of the ultracentrifugation tube.
7.5. Pour cell culture supernatant directly into the syringe barrel to fill the ultracentrifugation tube.
7.6. Top up the tube with PBS as necessary until full. NOTE: Allow the tube to slightly overflow if necessary, ensuring there are no air spaces in the ultracentrifugation tube and no air bubbles
7.7. Seal ultracentrifugation tube using a soldiering iron, ensuring the tube is airtight by squeezing the tube gently.
7.8. Place the ultracentrifugation tubes in the ultracentrifugation rotor, ensure the line marked at the top of the tube and the circle drawn at the base of the tube are facing outwards, where the EV pellet will form.
7.9. Ultracentrifuge for 2 hours at 120,000 x *g* at 4 °C.
7.10. Carefully remove the rotor from the ultracentrifuge.
7.11. Remove ultracentrifugation tubes from the rotor and place in the tube holder.
7.12. Attach a 16G needle to a 10 mL syringe.
7.13. Pierce the top of the ultracentrifugation tube and insert the needle 2 cm into the top of the tube and aspirate the supernatant into the syringe.
7.14. Decant this EV-depleted supernatant into a 1.5 mL tube and freeze at – 80 °C.
7.15. Reinsert needle into the tube and carefully aspirate the remaining supernatant and discard.
7.16. Cut the top of the tube with a pair of scissors.
7.17. Pour off the remaining supernatant in one quick action.
7.18. Allow the tube to hang upside down for 1 minute.
7.19. Pat dry any liquid that forms on the tube with paper towel.
7.20. Invert the tube and place in the tube holder.
7.21. Add 100 µL of PBS to the tube.
7.22. Using the tip of a pipette gently dislodge the EV pellet at the base of the tube using a circular motion in the area marked in step 7.1.
7.23. Vortex briefly (1-2 seconds), twice.
7.24. Label a new 13 mL ultracentrifugation tube by drawing a circle at the bottom of the tube, where the expected EV pellet with form and mark a line around the neck of tube for orientation in the ultracentrifugation tube rotor. Label tube with sample identifier.
7.25. Place tube in tube holder.
7.26. Add 12 mL PBS to a new ultracentrifugation tube using a clean syringe and needle.
7.27. Using a syringe and needle collect the 100 uL EV sample and add to the tube. Carefully mix the EVs and PBS and rinse the syringe and needle by gently collecting PBS and aspirating into the tube. NOTE: Avoid creating bubbles
7.28. Seal ultracentrifugation tube using a soldiering iron, ensuring the tube is air tight by squeezing the tube gently.
7.29. Place the ultracentrifugation tubes in the ultracentrifugation rotor, ensure the line marked at the top of the tube and the circle drawn at the base of the tube are facing outwards.
7.30. Ultracentrifuge for 1 hours at 120,000 x *g* at 4 °C.
7.31. Carefully remove the rotor from the ultracentrifuge.
7.32. Attach a 16G needle to a 12 mL syringe.
7.33. Pierce the top of the ultracentrifugation tube and insert the needle 2 cm into the top of the tube, aspirate the supernatant into the syringe and discard.
7.34. Cut the top of the tube with a pair of scissors.
7.35. Pour off the remaining supernatant in one quick action.
7.36. Allow the tube to hang upside down for 1 minute.
7.37. Pat dry any liquid that forms on the tube edge.
7.38. Place in the tube holder.
7.39. Add 100 µL PBS to the tube.
7.40. Using the tip of a pipette gently dislodge the EV pellet at the base of the tube using a circular motion in the area marked in step 7.24.
7.41. Vortex briefly (1 - 2 seconds), twice.
7.42. Pipette 100 µL PBS/EV solution in to a clean 1.5 mL tube and keep on wet ice. NOTE: EV are ready for downstream processing and may be frozen and stored at -80 °C.

### 8. Determination of EV size and concentration using Nanoparticle Tracking Analysis (NTA)

8.1. Preparing the system NOTE: A detailed method for the use of Nanoparticle Tracking Analysis (NTA) for determination of EV size and concentration was reported by Mehdiani *et al*. ^13^
8.1.1. Defrost samples and keep them at 4 °C.
8.1.2. Start the NTA software by clicking the software-icon.
8.1.3. The software will open in ‘cell check’ and prompt you to fill the flow cell with deionized water. Fill a 10 mL syringe with deionized water and push into the machine, ensuring air bubbles pass into the loading chamber.
8.1.4. Follow the on screen instructions to prepare the system through a quality check (QC).
8.1.5. The software will perform a cell check and give you a measure of ‘cell quality’. This should be very good to excellent.
8.1.6. Prepare the quality control consisting of a 100 nanometer polystyrene beads. Pipette 1 µL of the standard into 999 µL of deionized water (diluted stock). Subsequently, add 10 µL of the diluted stock to 2.5 mL of deionized water (QC sample). Mix the solution by gently vortexing for 2-3 seconds and by pipetting. NOTE: Quality control samples should be prepared freshly every day but the initial diluted stock sample is stable for 1 week at 4 °C.
8.1.7. Fill a 1 mL syringe with the 1 mL of the QC sample and remove all air bubbles from the syringe.
8.1.8. Inject without introducing air bubbles into the NTA sample loading chamber by gently tilting the tip of the syringe into the injection chamber, whilst simultaneously pushing the plunger. Inject up to 950 µL of the QC sample into the chamber.NOTE: Do not introduce air bubbles into the sample loading chamber.
8.1.9. The software will perform an auto alignment (focus the camera) and will check the voltage for Z-potential readings – the voltage graph should be a smooth U-shaped curve.NOTE: If the error message ‘voltage too low error’ the cell may be wet from cleaning or not secure or there may be an air bubble.
8.1.10. Once the auto alignment/voltage check is complete, use the camera position drop down to check all positions (0.1-0.9) for unusual marks – presence of these indicate the cell may need flushing and/or cleaning.
8.1.11. Following QC the system will display ‘ready for measurements’.
8.2. Determining EV size and concentration by Nanoparticle Tracking Analysis NOTE: A detailed description for EV quantification using NTA is described elsewhere ^13^.
8.2.1. Prime the flow cell with PBS. Open the ‘Pump/temp’ tab and under ‘pump’ click ‘run’ for pump 2 (PBS reservoir). This will run PBS through the cell for 1 minute then automatically stop. NOTE: You may see particles in the chamber but these can be cleared by pushing 10 mL of PBS through the loading port.
8.2.2. Fill a 10 mL syringe with PBS and load into the chamber with no air bubbles.
8.2.3. Create a protocol for the device to run that will measure multiple positions of the laser and average the particles per frame.
8.2.4. Check the particle count is less than 5 (as close to 0 as possible) before proceeding – flush with more PBS if necessary.
8.2.5. Prior to dilution, mix the EV sample by pipetting.
8.2.6. Dilute your sample 1:1000 in PBS and mix my pipetting. Fill a 1 mL syringe with your sample and load into the chamber with no air bubbles.
8.2.7. Check that the particle count is within the acceptable range (the bar above your count value should be in the green region, or close to it). If not, adjust your sample dilution accordingly. In between samples and sample dilutions load 10 mL PBS into the front of the machine to clear the chamber, then load 1 mL of your diluted sample. NOTE: Each sample will independent dilution based on the concentration of the sample. The particle count of the device should be between 50 - 200 particles per frame for accurate measurements.
8.2.8. Go to the ‘measurement’ tab in the software.
8.2.9. Click the ‘Run video acquisition’ button.
8.2.10. Enter a sample name and select a destination folder to save the measurement.
8.3. Create an EV standard operating procedure (SOP)
8.3.1. Create, Save and Load and EV SOP – that will measure particle number and size at a shutter speed of 100 and camera sensitivity of 80, 11 camera positions for 2 cycles – NOTE: unless your sample has special requirements, there is no need to change these settingsSOP.
8.3.2. Enter the dilution of your sample and add any other notes you wish to add.
8.3.3. Click ‘OK’ and the software will automatically start recording. NOTE: Do not touch the machine or nearby countertops during this time as vibrations will influence the final readings.
8.3.4. Once finished, the software will automatically load a pop-up table showing average particle counts and sizes for each camera position.
8.3.5. Camera positions with statistically unusual readings will automatically be excluded – if you would like to manually exclude any camera positions (or re-include excluded ones for any reason) click the check box to the left of the sample details.
8.3.6. Click ‘continue’ and the software will create and open a PDF of your sample and the size and concentration results.
8.4. Statistical Analysis
8.4.1. NTA data was analyzed by one-way or two-way ANOVA with post-hoc Tukey correction. Data are plotted as group means ± standard deviation. A P value of <0.05 was considered significant.

## REPRESENTATIVE RESULTS

We determined the quantity of EVs isolated from human gluteal adipocytes following the described protocol. We calculated the size and concentration of adipocyte-derived EVs using NTA (**Figure 1 A,B**). We utilized sham-media controls, which were equal volumes of media that had not been in contact with cells, but cultured and subject to the isolation procedure described above. We measured the adipocyte-derived EV concentration following the initial isolation and after washing the isolated adipocyte EVs in PBS (**Figure 1 A,B**) and plotted the group means ± standard deviation (SD), which was analyzed by a two-way ANOVA with post-hoc Tukey correction.

**Figure 1:**
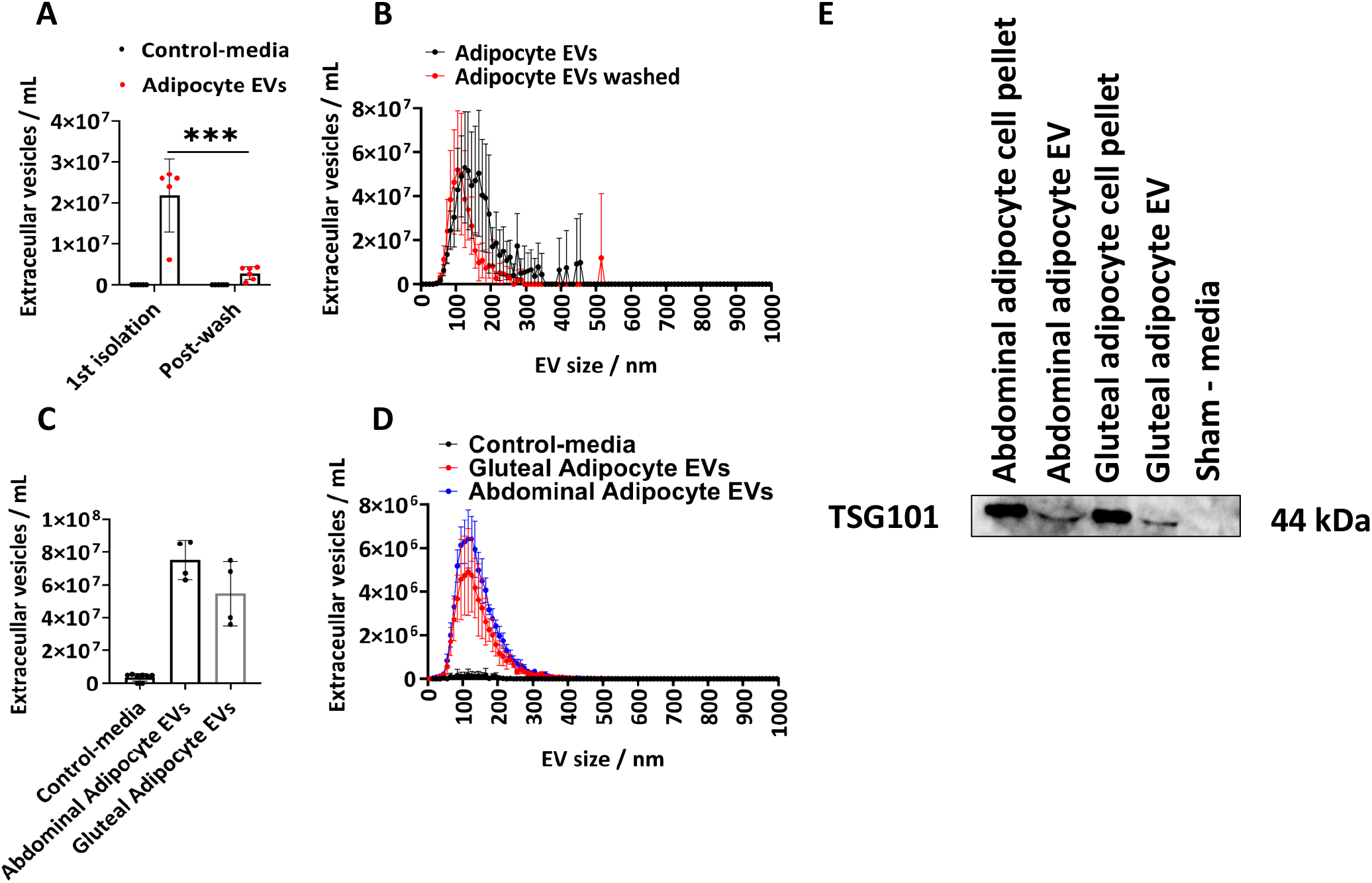
Size and distribution profile of adipocyte-derived EVs from cell culture media and determination of EV-protein TSG101. Total EV concentrations and size and concentration distribution profiles as determined by Nanoparticle Tracking Analysis (NTA) from the 1^st^ isolation (N=5) and following washing with PBS (N=6). (C) Total concentrations and (D) size and concentration distribution profiles determined by NTA for abdominal and gluteal derived EVs from T175cm2 flasks (N=4 per group). (E) Western blot of gluteal and abdominal derived EVs for TSG101. Cell pellets and sham-media were used as positive and negative controls respectively. Data are group means ± standard deviation (SD). A one-way or two-way ANOVA with post-hoc Tukey correction. *** P<0.001

The concentration of adipocyte-derived EVs determined by NTA from the 1^st^ isolation ranged from 6.10 x 10^6^ to 2.70 x 10^7^ with a median of 2.60 x 10^7^ EVs/mL (**Figure 1 A,B**). Following a PBS wash there were significantly fewer adipocytes derived EVs per sample (**Figure 1 A,B**) (P<0.001), which ranged from 5.00 x 10^5^ to 4.30 x 10^6^ with a median of 2.70 x 10^6^ EVs/mL. The sham- media controls contained no EVs as determined by NTA (**Figure 1 A,B**). The modal size of EVs from the first isolation was 125 nm and 105 nm following a PBS wash (**Figure 1 A,B**). The described protocol was further applied to abdominal and gluteal derived-adipocytes from larger T175 cm^2^ flasks. These gluteal EV samples derived from T175 cm^2^ flasks ranged in concentration from 3.60 x 10^7^ to 7.50 x 10^7^/mL with a median of 5.40 x 10^7^ EVs/mL. Abdominal adipocyte derived EVs from T175 cm^2^ flasks ranged in concentration from 6.30 x 10^7^ to 8.60 x 10^7^/mL with a median of 7.60 x 10^7^ EVs/mL (**Figure 1 C,D**). The modal size of EVs derived from the T175 cm^2^ flasks was 115 nm for gluteal EVs and 125 nm abdominal EVs. We confirmed the presence of EV-protein in gluteal and abdominal derived EVs by immunoblotting for tumor susceptibility gene 101 (TSG101) and found that abdominal and gluteal adipocyte cell pellets and abdominal and gluteal adipocyte-derived EVs were positive for TSG101, whereas as sham-control media, which had not been in contact with cell was negative (**Figure 1E**).

## DISCUSSION

We demonstrate a protocol for the isolation of gluteal and abdominal adipocyte-derived EVs from cell culture supernatants and determined their size and concentration by NTA^7,14,15^. We show that cultured human adipocytes produce and release EVs into the cell culture media, which can be subsequently isolated using filtration and ultracentrifugation. We determined the size and concentration profile of isolated adipocyte derived EVs and show that ultracentrifugation likely co-isolated contaminants from the cell culture media and that washing the isolated EV pellets in PBS significantly lowers their concentration in a second NTA measurement. We further determined the purity of the isolated gluteal and abdominal derived EVs by western blotting for TSG101 an EV-marker. Gluteal and abdominal derived EV preparations were positive for TSG101 but importantly this was absent in control-media that was not exposed to cells. The presented experiments used human adipocytes as the parent cell generating EVs but the described method is suitable for other cell types including, endothelial cells, vascular smooth muscle, skeletal muscle, immune cells and for the isolation of EVs from patient platelet poor plasma or serum.

Adipocyte derived EVs are elevated in metabolic diseases and determining alterations in the size and number of adipocyte EVs in vivo is complex because biological fluids such as blood contain EVs from a number of cell sources that are also implicated in the pathogenesis of metabolic disease, including EVs from endothelial cells, skeletal muscle, erythrocytes and immune cells. The method described here allows determination of human adipocyte EVs, which may provide a useful model for mechanistic studies investigating the factors that lead to EV biogenesis in adipocytes, which is currently unknown. Importantly, determining adipocyte EV biogenesis and how loading of particular RNAs, proteins and metabolites is orchestrated in adipocyte EVs may reveal novel therapeutic opportunities to perturb pathogenic adipocyte EV signaling in metabolic dysfunction. Detailed studies will provide a better understanding of how EV size, number, biogenesis pathway and EV-cargo (RNA, proteins and metabolites) are altered in response to disease or stimuli such as perturbations in oxygen, glucose, lipids and insulin. Information on the role of environmental factors on adipocyte EV signaling in metabolic disease and how adipocyte-derived EVs contribute towards adipose tissue inflammation may uncover novel therapeutic targets in metabolic disease.

### Limitations

#### in vitro generation of adipocyte derived EVs

The use of human preadipocytes in vitro provides a model system to study the release and generation of adipocyte-derived EVs following in vitro adipocyte differentiation but there are a number of limitations. In particular, in vitro derived adipocyte EVs are likely to differ from adipose-derived EVs retrieved from bio fluids, such as plasma ^14^ in their size, concentration, EV-protein,-RNA, –metabolites and function. These EV differences could be influenced by other non-adipocyte cells that are resident in adipose tissue in vivo, such as adipose tissue derived stem cells, endothelial cells and macrophages, which are intimately linked to adipose tissue physiology and have shown roles in adipose tissue pathology, including adipose tissue inflammation.

It should be noted that the two week in vitro differentiation protocol described here may not be sufficient to generate fully mature adipocytes equivalent to those seen in vivo; in vitro differentiated adipocytes grown in a two-dimensional format display a different morphology to in vivo cells and do not develop unilocular lipid droplets..Furthermore, the preadipocytes described in this protocol are obtained from the adipose stromal-vascular fraction and we have not assessed the contribution to EV pool from other cell types which were not completely eliminated during the cell isolation.

The loss of important cell to cell interactions of adipocytes with other non-adipocytes in adipose tissue may influence adipocyte EV generation, release,, EV-protein and EV-RNA from adipocytes and adipose tissue derived stem cells ^16^. However, an assessment of how in vitro derived adipocyte EVs differ from those produced in vivo has not been undertaken exhaustively.

Primary tissue biopsies contain blood and therefore the derived cell cultures may contain erythrocytes and erythrocyte derived EVs irrespective of the multiple washes and media changes highlighted in our protocol. An additional red blood cell lysis step following isolation of the stromal-vascular fraction may be necessary to eliminate the effects of erythrocytes on adipocytes. This is important because erythrocyte derived EVs can impact the cellular function of other cells ^17^ and erythrocyte derived EVs are elevated in the presence of oxidative stress ^18^ and in patients with metabolic syndrome ^19,20^. Therefore adipose tissue derived from metabolic disease patients may contain elevated levels of erythrocyte derived EVs, which may influence the in vitro phenotype of adipocytes.

#### Elimination of FBS

Our described protocol utilized FBS in the growth media during adipogenic differentiation but subsequently the adipocytes were subject to multiple media changes before the final media collection for the isolation of adipocyte derived EVs. Therefore, we assumed the overall risk for contamination of FBS derived EVs in ours EVs preparations to be low and subsequently confirmed that residual EVs were not present in the cell culture media by western blotting for TSG101. The isolation of cell cultured EVs from cell sources that require FBS must use EV-depleted FBS or deplete bovine-EVs through ultra-centrifugation to prevent bovine-EVs confounding adipocyte EV concentrations and analysis of adipocyte EV cargo. Depletion of serum from adipocytes is known to alter their cellular responses ^21^ and therefore investigation must ensure that serum depletion or EV-depletion from serum renders their adipocyte cultures truly representative of adipocyte biology.

#### Technical limitation of EV isolation using filtration and ultracentrifugation

We describe a method of ultracentrifugation with single use plastic tubes that require sealing prior to ultracentrifugation for EV isolations. We acknowledge that these single use sealed tubes may not be an economical option for many individuals and suggest exploration of similar tubes, which do not require sealing and are reusable. However, investigators must ensure that washing of reusable tubes is adequate and does not lead to progressive accumulation of protein, lipid and RNA contaminants overtime, which may impact downstream investigations of EV associated cargo or impact on cellular function studies.

The filtration and ultracentrifugation protocol described here has been used for numerous years and multiple studies have highlighted the short fallings of this method, including the non-specific isolations of contaminating cellular components such as cellular mitochondria, the presence of nuclear fragments and constituents of the cell membrane. Furthermore, the method described here will co-isolate lipoproteins produced by adipocytes or those present in EV-depleted FBS. The method here may be further developed by the use of density ultracentrifugation and size exclusion chromatography (SEC) to eliminate contaminating soluble proteins and some lipoproteins. Coupled with PBS washing of the isolated EVs and SEC the co-isolated contaminants can be limited but not completely eliminated. Therefore, users should ensure the inclusion of appropriate controls, including a sham-media that has not been in contact with cells to account for soluble proteins and lipoproteins in the culture media and an EV-depleted supernatant control to demonstrate the successful isolation of EVs from conditioned media that still contains soluble proteins and lipoproteins.

The isolation of EVs using filtration and ultracentrifugation relies on the operator ensuring undue pressure is not applied to the cell culture supernatants whilst it is passed through the bore of the filter or the bore of the needle/syringe. Application of undue force during this stage of the described protocol may rupture EVs, influence the final EVs concentrations and generate free RNA, proteins and metabolites, which were once encased in EVs. We have described a method here, which doesn’t require the syringe barrel and therefore the application of force to the EVs in conditioned media as they pass through the filter or needle into their collection reservoirs and ultracentrifugation tubes. Nonetheless, further care must be undertaken when resuspending the EV pellet in PBS. Only a brief vortex must be used, as vigorous vortexing may disrupt the EV membranes.

Following ultracentrifugation operators must be careful with the ultracentrifugation tubes as not to disturb the EV pellet. This can be achieved by handling the tubes carefully and moving them slowly between the ultracentrifugation rotor and the tube rack. Further care must be taken when piercing a hole in the top of ultracentrifugation tube to aspirate the EV-depleted supernatants. Inserting the needle into the top of the tube and aspirating the supernatants quickly will create a vacuum in the syringe barrel, which can violently force the supernatant back into the ultracentrifugation tube and disrupt the EV pellet. After cutting the ultracentrifugation tube to pour off the remaining supernatant, care must be taken because the EV pellet may become loose and pouring will discard the EV pellet. Alternatively, a syringe and needle can be used to slowly remove the remaining supernatant without pouring or inverting the tube.

Filtration and ultracentrifugation of supernatants for the isolation of EVs is a useful and efficient method. But it is liable to the co-isolation of lipoproteins and soluble proteins. These can be mitigated by washing the EVs with PBS (as described) but this will not eliminate all contaminants. Soluble proteins can be eluted from the EV fraction by utilizing SEC), however this method does not distinguish between lipoproteins and EVs. SEC-elutions that contains EVs can be combined with ultracentrifugation to pellet EVs. Filtration and differential ultracentrifugation is a preferred method of EV isolation over precipitation techniques, which use polyethylene glycol because these precipitation methods co-isolate large quantities of soluble proteins and lipoproteins in cell culture supernatants and other biological fluids. Ultracentrifugation is likely to remain the most accessible form of EV isolation for many because most laboratories are equipped with an ultracentrifuge, therefore mitigating initial startup costs. But for many ultracentrifugation for EV isolation is hindered by the volume of ultracentrifugation tubes and the starting volume of material. Several 100 mL’s of culture supernatants may be needed to produce sufficient EV quantity for downstream proteomics or RNA-sequencing. However, it is likely that ultra-centrifugation techniques for EV isolation will be accompanied by other techniques such as SEC and immuno-affinity capture using tetraspanins CD9, CD63 and CD81 to improve the purity of isolated EVs. Other techniques such as commercially available precipitation solutions and flow cytometry may be of some use in specific investigations.

#### Purity of EV preparations

We confirmed the isolation of adipocyte derived EVs by western blotting for TSG101 but this single western blot falls short of the guidelines published by the international society for extracellular vesicles (ISEV). Further characterization of these adipocyte derived EVs would be ideal using tetraspanins CD9, CD63 and CD81 to identify exosomes and markers of cellular contamination such as histone H3, albumin and apolipoprotein A1.

The protocol presented here allows the isolation of EVs from cell culture supernatants from a range of cell sources including adipocytes for determination of EV size, concentration, EV-markers by western blot and utility in omics based technologies such as proteomics and RNA- sequencing.

## Data Availability

n/a

## ACKNOWLEDGMENTS

N.A. and R.C. acknowledge support by research grants from the British Heart Foundation the Centre of Research Excellence, Oxford (N.A. and R.C.; RE/13/1/30181 and RE/18/3/34214), British Heart Foundation Project Grant (N.A. and R.C.; PG/18/53/33895), the Tripartite Immunometabolism Consortium, Novo Nordisk Foundation (NNF15CC0018486), the National Institute for Health Research (NIHR) Oxford Biomedical Research Centre (BRC), Nuffield Benefaction for Medicine and the Wellcome Institutional Strategic Support Fund (ISSF). The views expressed are those of the author(s) and not necessarily those of the NHS, the NIHR or the Department of Health.

